# Comparing a trial of labour with planned caesarean after one previous caesarean delivery: a protocol for a population-based cohort study

**DOI:** 10.64898/2026.07.12.26357849

**Authors:** Romy Ehrlich, Pejman Adily, Mark Lauer, Rajit Narayan, Hala Phipps, Adam Mackie, Vincenzo Berghella, Adrienne Gordon, Bradley de Vries

## Abstract

The caesarean section rate has risen. An increasing number of women who have had one previous caesarean section need to consider mode of delivery in a subsequent pregnancy. Both trial of labour after caesarean (TOLAC) and planned repeat caesarean section are associated with maternal and perinatal risks. We will undertake a retrospective cohort study of recorded livebirths for women who have had one previous caesarean section in the United States from 2011 to 2024. Using multivariable logistic regression, we will examine the relationship between the intended mode of delivery at 39 weeks gestational age (TOLAC at 39 weeks or any mode of delivery at 39 to 43 weeks versus caesarean section without trial of labour at 39 weeks) and important maternal and neonatal outcomes. We will also examine the relationship between TOLAC and caesarean section without a trial of labour for maternal outcomes at 20 to 43 weeks and for perinatal outcomes at 37 to 43 weeks. Due to the large data size, this study will be able to report on rare maternal and perinatal outcomes and contribute to evidence that influences important decisions about mode of delivery after one caesarean section.

## Background

Caesarean section (CS) rates have risen substantially in many countries over recent decades [1]. Now, approximately one third of all women give birth via CS in Australia (41% in 2023) [2], the United States (US) (32.4% in 2024) [3] and the United Kingdom (37.8% in 2023) [4]. In many settings, a previous CS is now the single most common indication for a subsequent CS [5]. In 2023 in Australia, only 11% of women with one previous CS birthed their second baby vaginally [2]. Consequently, counselling and management of the next birth after CS is a frequent and clinically important component of maternity care.

When planning mode of delivery, the risks of a planned CS must be balanced against the risks of a trial of labour after caesarean section (TOLAC). Beyond the short-term risks associated with planned CS such as bleeding, infection, venous thromboembolism and visceral injury, serious long-term maternal morbidity related to placenta praevia, placenta accreta spectrum and peripartum hysterectomy increases in a dose-response manner with each additional CS [6]. The primary concern regarding TOLAC is the risk of uterine rupture, typically considered to be approximately 1 in 200 [7], with potentially serious consequences including perinatal and maternal mortality, neonatal hypoxic ischaemic encephalopathy, and maternal hysterectomy [8–12]. An additional uncommon but critical risk of TOLAC is the prospective risk of antepartum stillbirth whilst awaiting spontaneous labour [7].

Maternity care providers often discuss these risks with women during antenatal visits in the third trimester, with the goal of establishing a plan for CS at 39 weeks or an intention to undergo a trial of labour following either spontaneous onset or induction of labour at 39 weeks or beyond. However, intervening maternal or fetal conditions such as spontaneous preterm labour, pre-eclampsia or fetal growth restriction may require a decision regarding mode of delivery be made at any gestation, including during the preterm or early term period.

There is a lack of robust, randomised trial evidence to guide clinicians and women in their decision-making regarding mode of delivery during antenatal counselling. The largest and highest quality evidence to date regarding TOLAC versus planned CS are Landon’s 2004 landmark observational study [8] and Guise’s subsequent systematic review in 2010 [12]. However, neither were powered to comment on certain critical though rare maternal and neonatal outcomes. Hence, high-quality comparative data that incorporates modern obstetric practice, stratifies by gestational age, and is adequately powered for rare but serious outcomes is required to inform decision-making.

## Description of Protocol

### Objectives

This study aims to use a large United States population-based dataset to compare the maternal and perinatal outcomes of a TOLAC versus a CS without trial of labour in women who have had one previous CS and no previous vaginal births.

We aim to answer two clinical questions.

1. Among women with one previous CS, how do the maternal and perinatal outcomes differ between (i) a planned elective repeat CS at 39 weeks of gestation and (ii) an intention to undergo TOLAC at 39 weeks (following spontaneous labour or induction of labour)
2. Among woman with one previous CS, how do the maternal and perinatal outcomes differ between CS without a trial of labour versus a trial of labour at each gestational age between 20 and 43 weeks?

### Study Design

This will be a retrospective cohort study examining the relationship between mode of delivery (TOLAC or caesarean section without TOLAC) and important maternal and neonatal outcomes. This study will be reported according to STROBE Guidelines [13].

### Study Setting

All eligible births recorded in the USA using the 2003 Revision of the US Certificate of Live Birth between 2011 and the most recent year for which data are available (currently 2024). Ten to 14% of data for the years 2011 to 2014 do not use the 2003 Revision and will be excluded. For the outcome of stillbirth or perinatal death, period-linked and stillbirth data will be used where data are available (currently 2014 to 2023).

### Participants

#### Eligibility criteria

- 20 to 43 weeks gestational age (inclusive)
- Exactly one previous live birth
- Exactly one previous CS
- The 2003 Revision of the US Certificate of Live Birth was used

#### Exclusion criteria

- Fetal congenital anomaly where reported (anencephaly, meningomyelocele/spina bifida, cyanotic congenital heart disease, congenital diaphragmatic hernia, omphalocele, gastroschisis, limb reduction defect, cleft lip/palate, Down Syndrome, suspected chromosomal disorder, hypospadias)
- Multiple pregnancy
- Non-cephalic presentation

### Variables

#### Primary Study Factor

Whether there is a TOLAC or a CS without a trial of labour.

#### Other Variables

- Maternal age (years)
- Maternal pre-pregnancy body mass index (kg/m^2^)
- Maternal height (cm)
- Maternal race (black, white, indigenous American or Alaskan Native, other)
- Maternal educational attainment (12^th^ grade or less, high school graduate or GED/some college credit/associate degree, Bachelor’s degree, Master’s degrees, PhD or Professional degree)
- Pre-pregnancy cigarette smoking (yes/no)
- Payment type (public or private)
- Trimester of onset of maternity care (four categories: first, second, third, no antenatal care)
- Year of delivery (categorical)
- Gestational age at birth (weeks, categorised)
- Birthweight (grams)
- Birthweight centiles (neonatal sex and gestational age adjusted)
- Inter-pregnancy interval (months)
- Labour type (spontaneous v induced/augmented)
- Diabetes in pregnancy (pre-existing, gestational, none)
- Hypertension in pregnancy (yes/no)

Mean and standard deviation (SD) will be used to describe continuous variables if they are normally distributed. Otherwise, median, and interquartile range (IQR) will be used. Numbers and percentages will be used for categorical variables.

### Data Sources / Measurement

Publicly available data from the Center for Disease Control (CDC) will be used and can be downloaded from https://www.cdc.gov/nchs/data_access/vitalstatsonline.htm.

Variables that are a planned part of the analysis will be interrogated to describe any missingness and extreme outliers.

### Outcomes

These are all binary outcomes and will be expressed as numbers and percentages (n %)

Maternal outcomes:

- Ruptured uterus
- Unplanned hysterectomy
- Blood transfusion
- Admission to intensive care unit (ICU)

Perinatal outcomes:

- Stillbirth or neonatal death
- Apgar score at 5 minutes < 6
- Neonatal seizure or serious neurological dysfunction (severe alteration of alertness excluding lethargy or hypotonia in the absence of other neurological findings)
- Admission to neonatal intensive care unit
- Neonatal ventilation immediately after birth
- Neonatal ventilation for more than 6 hours

### Sample Size

No sample size calculation will be performed. The maximum amount of available data will be used.

### Statistical Methods

Data will be downloaded from the CDC website (https://www.cdc.gov/nchs/data_access/vitalstatsonline.htm) for birth years 2011 to 2024.

The comparisons will be:

1. Between TOLAC and planned CS (without TOLAC)
2. Between TOLAC without induction or augmentation of labour, TOLAC with induction or augmentation of labour, and birth by CS without TOLAC

TOLAC includes caesarean deliveries where a TOLAC was attempted, and all vaginal births.

The analysis will compare TOLAC with CS as follows:

#### (1) Perinatal and maternal outcomes at 39 to 43 weeks gestational age

As a decision for a TOLAC or no TOLAC is often made by 39 weeks gestational age, we will compare outcomes for women and their babies (i) who had a TOLAC at 39 weeks gestational age or any births at 40 weeks or more whether or not a TOLAC occurred with; (ii) those who had a planned CS at 39 weeks gestational age (i.e., the primary study variable will be dichotomous). Planned CS will be defined as all CS where a TOLAC was not attempted. This approach is designed to approximate an intention-to-treat analysis and assumes that if a planned CS was not performed at 39 weeks, then the intention *at this time* was for a TOLAC. The results are expected to inform a decision for a TOLAC versus planned CS at 39 weeks, accounting for the possibility that an indication for CS might arise later in the pregnancy.

#### (2a) Perinatal outcomes for births at 37-43 weeks gestational age

The analysis for perinatal outcomes will be limited to term births (37 weeks or more) because preterm planned CS are often performed for serious concerns about fetal wellbeing. This could bias any comparison between the TOLAC and planned CS groups, potentially favouring the TOLAC group. However, we cannot predict the direction of any bias with certainty.

Perinatal outcomes following a TOLAC will be compared with perinatal outcomes following a CS without TOLAC. The study variable will have three categories: (i) TOLAC with no induction or augmentation of labour; (ii) TOLAC with induction or augmentation of labour; (iii) CS without TOLAC. Based on outcomes for the baby, the results are expected to inform the decision between a CS and a TOLAC for women in spontaneous labour at 37-43 weeks gestational age; and to inform the decision between a CS and an induction of labour when delivery is required and labour has not yet commenced.

#### (2b) Maternal outcomes for births at 20-43 weeks gestational age

The study variable will have three categories: (i) TOLAC with no induction or augmentation of labour; (ii) TOLAC with induction or augmentation of labour; (iii) CS without TOLAC. Based on maternal outcomes, the results are expected to inform decisions between a TOLAC and planned CS for women in spontaneous labour from 20 to 43 weeks. They are also expected to inform the decision between induction of labour and planned CS when delivery is required, and labour has not commenced.

#### (3) Sensitivity analysis

Subgroup analyses will be performed for maternal outcomes from 20-43 weeks gestational age within the following gestational age groups:

i. 20-29 weeks
ii. 30-36 weeks
iii. 37-38 weeks
iv. 39-43 weeks

The subgroup analyses will use the variables in the final regression model (after backward stepwise regression – see Analysis section below) for Analysis 2b in Table 1. These analyses will allow a qualitative comparison of observed relationships between mode of delivery and adverse maternal outcomes in different gestational age groups.

**Table 1.** Summary of planned analyses.

| Analysis | Population | Comparison | Outcomes | Potential value |
| --- | --- | --- | --- | --- |
| 1 | 39 or more weeks gestational age | (i) TOLAC at 39 weeks OR birth by any means at 40 to 43 weeks; versus (ii) Planned CS at 39 weeks (two groups)* | Perinatal and maternal | Aid in decision making at term for TOLAC versus planned CS (approximate intention-to-treat analysis) |
| 2a | 37 to 43 weeks gestational age | (i) TOLAC without augmentation or induction; versus (ii) TOLAC with augmentation or induction; versus (iii) CS without TOLAC (three groups)** | Perinatal | Aid in delivery-planning for women in spontaneous labour, or who require early term delivery |
| 2b | 20 to 43 weeks gestational age | (i) TOLAC without augmentation or induction; versus (ii) TOLAC with augmentation or induction; versus (iii) CS without TOLAC (three groups) | Maternal | Aid in delivery-planning for women in spontaneous labour, or who require delivery |
| 3 | Sensitivity analyses | Analysis 2b will be replicated within gestational age groups (i) 20-29 weeks; (ii) 30-36 weeks; (iii) 37-38 weeks; (iv) 39-43 weeks | Maternal | Assess for differences in the relationship between type of birth*** and maternal outcomes by gestational age group |

\* This approach assumes that if a planned CS was not performed at 39 weeks gestational age, then the intention *at this time* was for a TOLAC. The purpose is to simulate an intention to treat analysis.
\*\* As the stillbirth data do not specify if labour was spontaneous, augmented or induced, for the outcome of perinatal death, TOLAC will be compared with CS without TOLAC.
\*\*\* Type of birth refers to (i) TOLAC without augmentation or induction; (ii) TOLAC with augmentation or induction; or (iii) CS without TOLAC

#### Analysis

For the 2023 subset of data, we conducted preliminary analyses to inform modelling decisions. We plotted quantile groups of each continuous variable against the log-odds of each outcome for both TOLAC and planned CS at 39 weeks (Analysis 1), to assess for potential interaction with TOLAC versus planned CS and to assess for linearity. We detected no interactions between explanatory variables and TOLAC versus planned CS for any of the outcomes. For variables with no evidence of non-linearity, we will treat variables as continuous. Non-linear variables will be treated as described in the list of variables below. Gestational age is expected to have a strong non-linear association with many outcomes and will be treated as categorical. The same model specification was re-used for Analysis 2.

All outcomes are binary and will be analysed using multivariable logistic regression adjusting for the potential confounders listed below:

- Gestational age in weeks:
  ∘ Analysis 2a (Table 1): 7 categories (37, 38, 39, 40, 41, 42, and 43 weeks)
  ∘ Analysis 2b (Table 1): 4 categories (20-29, 30-36, 37-38, and 39-43 weeks)

- Maternal age in years (Linear for all outcomes except maternal blood transfusion. For maternal blood transfusion a maternal age-squared term will be included in the base model)
- Pre-pregnancy maternal body mass index (BMI) (Continuous for BMI ≥ 25; If BMI < 25 kgm^-2^ then BMI will be set to 25 kgm^-2^)
- Interpregnancy interval - the time from the previous CS to the onset of the current pregnancy (Linear spline with one knot at 21 months) [14]
- Maternal height in cm (Linear)
- Birthweight centile for perinatal outcomes (Categorical in three groups: (i) lowest 5 centiles; (ii) middle 90 centiles; (iii) and highest 5 centiles). Centiles will be calculated using published birthweight standards. [15]
- Raw birthweight for maternal outcomes
- Birth year (Categorical by year)
- Race (Categorical: white; black; American Indigenous or Alaskan Native; or Other)
- Cigarette smoking in pregnancy (Dichotomous)
- Payment type (Dichotomous: public/private)
- Mother’s Education (Five categories: (i) 12th grade or less; (ii) high school graduate or GED, some college credit, or associate degree; (iii) Bachelor’s degree; (iv) Master’s degree; (v) PhD or Professional degree)
- Hypertension in pregnancy (Dichotomous)
- Diabetes (Categorical: pre-pregnancy, gestational, or no diabetes)

If data missingness is present for the explanatory variables for more than 5% and less than 40% of eligible births, then multiple imputation will be considered. Potential mechanisms of missingness and the relationship of missingness to the outcome and covariates will be assessed when deciding to conduct multiple imputation or a complete case analysis.

If multiple imputation is required, we will use fully conditional specification, with five imputed datasets. Any multiple imputation will include all explanatory and outcome variables, any spline terms, and derived terms including BMI, and any squared terms used to assess for linearity. Interdependencies amongst variables will be handled with the passive imputation method.

Where required, the linearity of continuous explanatory variables will be tested by adding a squared term for the continuous variable. Based on our preliminary analysis, the only squared term will be maternal age for the outcome of blood transfusion.

Backward stepwise elimination will be conducted using log-likelihood ratio tests on the contribution of each explanatory variable remaining in the model. Variables will be eliminated one at a time if (1) they have the highest p-value; and (2) the p-value is > 0.05. If multiple imputation has been used, p-values for the likelihood ratio test in each imputed dataset will be pooled using the median p-value rule.

### Bias

Possible sources of bias have been considered including:

1. The data do not record planned TOLAC versus planned CS, and a direct intention-to-treat analysis is therefore not possible. We will approximate an intention-to-treat analysis by comparing two groups:
  a. No planned TOLAC at 39 weeks: Birth by planned CS at 39 weeks with no trial of labour.
  b. Planned TOLAC at 39 weeks: Vaginal birth at 39 weeks, or a CS with a trial of labour at 39 weeks; AND any birth beyond 39 weeks (including CS without TOLAC). The latter assumes that any woman who reached 40 weeks or more did not plan a CS without labour at 39 weeks. We expect that some participants in (a) No planned TOLAC at 39 weeks will have chosen a TOLAC but had a CS for clinical indications, and conversely, some participants in (b) Planned TOLAC at 39 weeks will have chosen a CS but may have had a vaginal birth for example due to a rapid labour. We expect these to constitute a small fraction of the population and that they will not substantially bias the results.
2. Planned preterm CS are often performed for concerns about fetal wellbeing. We therefore considered the risk of bias for perinatal outcomes at preterm gestational ages to be very high and did not assess perinatal outcomes at gestational ages < 37 weeks. Bias is also possible for the maternal outcomes such as blood transfusion. For example, someone with a CS for placenta praevia will have a higher risk of a blood transfusion and will be in the CS group, potentially biasing the results in favour of TOLAC. We expect that this will occur in only a small proportion of participants and that any bias will be small.
3. Maternal and perinatal outcomes may be influenced by covariates such as maternal age, cigarette smoking, and maternal diabetes. We have adjusted for all available variables that we considered could act as confounders.

### Summary

This study will utilise 14 years of all recorded US birth data to compare maternal and perinatal outcomes for women who have had one previous CS undergoing TOLAC with repeat CS without trial of labour. Due to the size of the dataset, we will be uniquely able to comment on rare maternal and perinatal outcomes. This will provide an up-to-date contribution to the growing body of evidence to aid clinical decision-making and counselling around TOLAC at any gestation of pregnancy.

## Protocol validation

This study will be reported in accordance with the STROBE guidelines [16]. To minimise the risk of overfitting, a preliminary analysis of the 2023 subset of the data was undertaken to prespecify the modelling strategy, including variable categorisation, assessment of linearity, and consideration of clinically plausible interaction terms. The 2022 subset was used for the stillbirth or neonatal death outcome due to data availability. The final model will be developed in the complete study dataset according to this prespecified strategy.

We conducted preliminary analyses using the 2023 subset of the US data (except for stillbirths and neonatal deaths for which we used the 2022 data) to inform modelling decisions. To assess for potential interaction with TOLAC versus planned CS and to assess for linearity, we plotted quantile groups of each continuous variable against the log-odds of each outcome for both TOLAC and planned CS at 39 weeks. We detected no interactions between explanatory variables and TOLAC versus planned CS for any of the outcomes. The linearity of variables and the subsequent model specifications for each analysis is outlined above.

## Limitations

For large datasets with many contributors, data entry errors are expected, and some data fields are not available. Uterine rupture is defined in the data as a full-thickness disruption of the uterine wall involving the overlaying visceral peritoneum. Cases where the fetus, placenta and umbilical cord remained contained within the uterine cavity and silent or incomplete ruptures or asymptomatic separations were excluded. As this represents the more severe end of the spectrum of uterine disruption, rates of uterine rupture may be underestimated. However, data entry errors and any underestimation of uterine rupture rates are likely to be non-differential, occurring in both TOLAC and non-TOLAC groups, and are not expected to influence the relative risks of adverse outcomes. Additionally, information may be limited or lacking regarding some important confounders, such as detailed obstetric history beyond parity, indication for prior CS, provider and hospital characteristics.

## Data Availability

All data produced in the present study are available upon reasonable request to the authors.

## Acknowledgements

We would like the thank Dr Tanis Fenton from the University of Calgary who provided details from the third-generation Preterm Growth (‘Fenton’) Charts that allowed us to calculate birth weight centiles for this study.

This research did not receive any specific grant from funding agencies in the public, commercial, or not-for-profit sectors.

